# MAJOR AETIOLOGIES OF MALE INFERTILITY AMONG COUPLES ATTENDING FERTILITY CLINICS IN OSUN STATE, NIGERIA: FINDINGS FROM A MIXED METHOD STUDY

**DOI:** 10.1101/2024.08.10.24311725

**Authors:** Kehinde Awodele, Sunday Charles Adeyemo, Eniola Dorcas Olabode, Adeniyi Olaonipekun Fasanu, Akintunde Rasaq Akindele, Olusegun Adegboyega Afolabi, Olufunso Abidemi Olagunju, Olusegun Oyerinde, Lanre Olaitan

**Affiliations:** Obstetrics and Gynecology Department, Uniosun Teaching Hospital, Osogbo, Nigeria; Institut Superieur de Sante, Niamey, Niger Republic; Department of Community Health, Obafemi Awolowo University, Ile-Ife, Nigeria; Department of Obstetrics and Gynecology, Osun State University, Osogbo, Nigeria; Department of public health, University of Ilorin; Department of Pediatrics, Osun State University, Osogbo, Osun State, Nigeria; Faculty of basic medicine science Adeleke University, Ede

**Keywords:** Male Infertility, Implications, Seminal fluid Analysis

## Abstract

Male infertility accounts for nearly half of the infertility cases globally. Seminal fluid analysis (SFA) is a critical diagnostic tool in the evaluation of male infertility. This study aimed to assess the implications of seminal fluid analysis on male infertility among patients attending fertility clinics in Osogbo, Nigeria.

The study employed mixed-method approach of both qualitative (Key informant interview) among 10 participants and quantitative method (cross-sectional survey) using pre-tested structured questionnaire among 305 respondents. The respondents in the cross-sectional survey were also made to undergo seminal fluid analysis. The data from the qualitative study was analysed using Atlas ti while data from the quantitative study were analysed using IBM Statistical Product for Service Solution (SPSS) version 27. 0. Descriptive statistics was carried out for all variables. The univariate, bivariate and multivariate analysis were done using p<0.05 as level of significance.

The seminal fluid analysis of the respondents revealed that 241 (79.0%) had Normal sperm count (>32 million per ejaculation) while 64 (21.0%) had abnormal sperm count. Only 101 (33.1%) had normal progressive motility (>32 percent) while 204 (66.9%) had abnormal (Athenospermia) progressive motility. 195 (63.9%) were found to have abnormal morphology (Teratospermia i.e., <4%). The qualitative analysis further analysed the implications of SFA parameters on infertile males and these were substantial, extending beyond physical health to encompass psychological, emotional, and social well-being.

The study concluded that lifestyle modifications and early diagnosis as well as prompt treatment of medical conditions can curb high prevalence abnormality of SFA, hence reduce male infertility in our environment. The study recommends that advocacy program, early screening and public health education will further reduce the burden of infertility among the female folks.

## Introduction

Seminal fluid plays a crucial role in the fertilisation process, providing a supportive environment for sperm transport, nourishment, and protection against the female reproductive tract’s hostile conditions. [1] Seminal fluid analysis (SFA) is a critical diagnostic tool in the evaluation of male infertility. It involves the examination of various semen parameters, such as sperm concentration, motility, morphology, and seminal plasma composition, to assess a man’s fertility potential. [2] Male infertility accounts for nearly half of the infertility cases globally. In Southwest Nigeria, there is limited research on the seminal fluid analysis and its implications on male infertility among patients attending fertility clinics. Investigating this relationship can help identify factors contributing to infertility in this region and inform the development of targeted interventions to improve reproductive outcomes for affected couples. Understanding the connection between SFA findings and male infertility could have significant implications for clinical practice, public health policies, and patient counselling. Therefore, this study aimed to assess the implications of seminal fluid analysis on male infertility among patients attending fertility clinics in Osogbo, Nigeria.

## Materials and Method

The study employed mixed-method approach of both qualitative (Key informant interview) among 10 participants and quantitative method (cross-sectional survey) among 350 respondents. The study population were male patients attending fertility centers in Osogbo, Osun State. Fisher’s formula (n=z^2^pq/d^2^) was used to determine the sample size. The respondents were selected using multistage sampling technique. 5 centers were randomly selected out of the major fertility centers in Osogbo. Proportional allocation was used to determine the number of respondents from each center. Male patients aged 18-50 were randomly selected in each center. Data was collected from the respondents of the quantitative study using pre-tested questionnaire as well as seminal fluid analysis. After consent has been obtained, the semen of the respondents was collected under sterile conditions after an abstinence period of 2–7 days using sterilized universal bottles. This was carried out in a private room near the laboratory in order to reduce the exposure of the semen to fluctuations in temperature and to control the time between collection and analysis. The semen was then analysed in the laboratory for sperm count, motility and morphology of the semen. The data from the questionnaire were analysed using IBM Statistical Product for Service Solution (SPSS) version 27. Descriptive statistics was used for all variables. Bivariate and multivariate analysis were done at p<0.05 as level of significance. Data from the key informant interview was recorded, transcribed and analysed using Atlas ti.

## Results and Discussion

### Results

#### Quantitative study

##### Sociodemographic characteristics

All the respondents were married male attending the infertility clinic in Osogbo. More than half 204(58.3%) of the respondents were with the ages of 30-45years, while the majority, 333 (95.4%) were Yoruba. 222 (63.6%) had tertiary level of education. More than half, 207 (59.0%) were skilled workers.

##### Seminal fluid analysis of male patients attending fertility center in

Majority of the respondents, 249 (71.0%) had Normal sperm count (>32 million per ejaculation) while 101 (29.0%) had abnormal sperm count in different categories. (Figure 1) Only 116 (33.1%) had normal progressive motility (>32 percent) while 234 (66.9%) had abnormal (Athenospermia) progressive motility. 224 (63.9%) were found to have abnormal morphology (Teratospermia i.e., <4%). (Table 1) Overall, 321 (91.8%) of the respondents have at least one abnormality in the semen analysis.

**Table 1:** Seminal fluid analysis of male patients attending fertility centers in Osogbo, Osun State.

| Variables | Frequency (n=305) | Percentage |
| --- | --- | --- |
| **Sperm Count** |  |  |
| Abnormal (>32 million) | 88 | 28.9 |
| Normal (<32 million) | 217 | 71.1 |
| **Progressive Motility** |  |  |
| Normal (> 32%) | 101 | 33.1 |
| Abnormal/Athenospermia (<32%) | 204 | 69.9 |
| **Morphology** |  |  |
| Normal (>4%) | 110 | 36.1 |
| Abnormal/Teratopsermia (<4%) | 195 | 63.9 |
| **Culture of any organism** |  |  |
| No | 305 | 100.0 |

**Figure 1:**
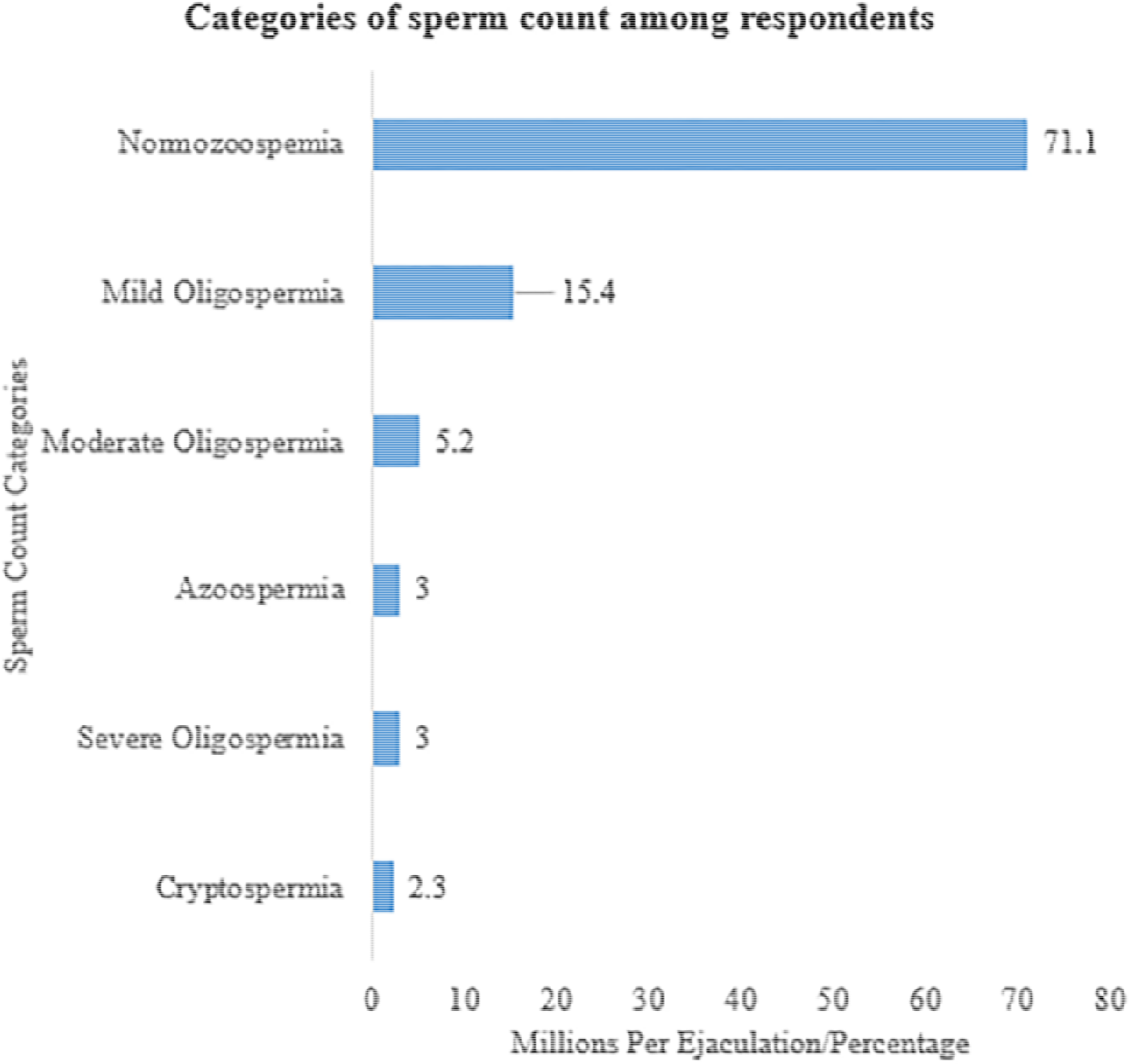
Categories of sperm count of male patients attending fertility centers in Osogbo, Osun State.

##### Qualitative study

##### Sociodemographic characteristics of participants

The participants’ ages range from 25 to 44 years old. Six participants were female, and four were male. Seven participants were married, three were single. The predominant ethnicity was Yoruba, with one participant being Nupe. Five participants were Christian and five were Muslim. Educational status varies, with three participants having tertiary education, two with HND, one with SSCE, and four with an unspecified level of education. The participant types include three wives, four nurses, and three doctors. (Table 2)

**Table 2:**
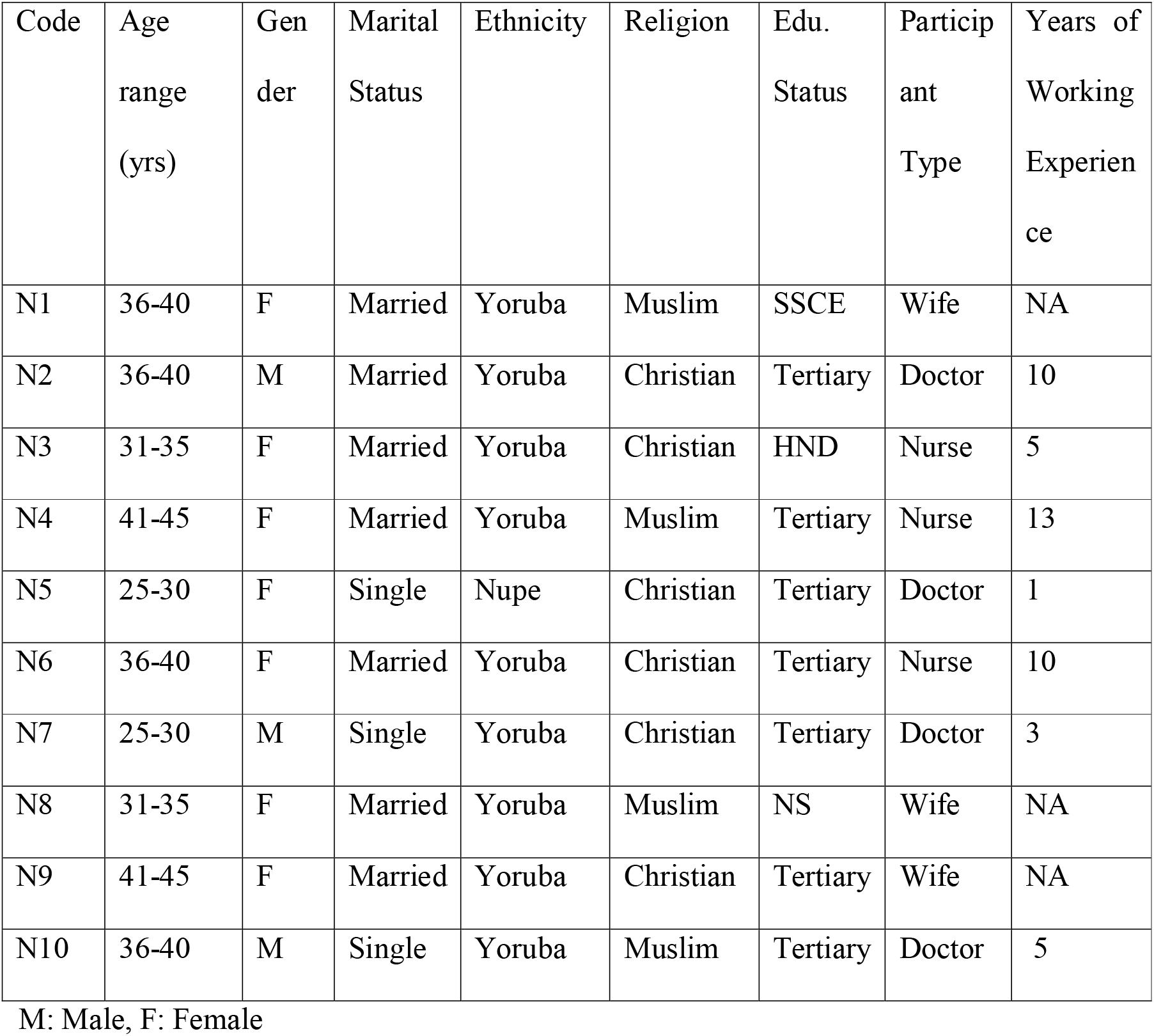
Sociodemographic characteristics of participants in the Key informant interview.

##### Causes of male infertility

Participants have different response what causes infertility. Majority of the response were directly or indirectly referring to the lifestyle such as smoking, alcohol consumption, having multiple sexual partners etc. as the cause of infertility, while some stated infertility is caused by medical conditions, genetics or childhood factor.

“*I think the lifestyle is one of the main causes, like smoking, taking alcohol, having multiple sexual partners which can predispose them to sexually transmitted infections” …. N1. “I think um the causes are diabetes, hypertension as well as obesity there are also risk factors like smoking and alcoholism… and some other dangerous lifestyles like having multiple sexual partners” N2. “I would say maybe smoking and alcohol” … N7. “It can cause by maybe disease”* … *N5*.

*“Well, it could be due to infections. Like we were told that in childhood, especially maybe a male child that was infected with mumps. If the mumps is not well treated, it could lead to infertility in future. And, it could be due to sexually transmitted infections that are not well*

*treated” … N4, …. This may be the one contracted during the bad habit of promiscuity by ether the male or female… and thereby causing infertility” …. N8*

*“Well, I think there are different causes of infertility: Ranging from genetics to lifestyle could affect the patient” …N6. It can be genetically motivated or it medical related causes…. N10*

##### Lifestyle as a cause of male infertility

All the participants affirmed that lifestyle could cause infertility in male. Factors such as alcohol consumption, smoking, multiple sexual partners, and untreated infections were stated as the lifestyles causing male infertility.

*“Yes, I think lifestyle. like a male that is having so many sexual partners could lead to infections, and if it’s not treated, it could lead to infertility” … N4. “Yes, I think yes, social life is causing it. Social life that can bring a lot of infections and some infections can cause infertility in men” … N5. “Smoking… Alcohol intake*…*N6, … “yes like engagement in drinking of alcohol, smoking etc*.*…N7. “Most of the times, this infertility is caused by lifestyle factors including smoking, alcohol consumption…. N10*.

##### Medical illness causing male infertility

While the majority of the participants affirmed that hypertension and diabetes may cause infertility, a few stated it can be caused by sexually transmitted infections.

**“***I know of diabetes, hypertension, but I’m not sure I have heard obesity being a cause of male infertility…N5. …Obesity…N2, I think especially sexually transmitted infections can cause it…N4. “Some underlying medical conditions such as diabetes, varicocele, that could also cause infertility” …N7 “I really can’t say” …N1. I heard the infections from usually from extra-marital affairs causes it… N9*.

## Discussion

This study assessed the major aetiology of infertility among couples attending fertility clinics in Osogbo, Osun State, Nigeria. More than half of the respondents were between the ages of 30 and 45 years. More than half of the respondents have tertiary as their highest educational attainment and the same proportion were skilled workers. This is similar to another study conducted among male partners attending infertility clinic in a Tertiary Nigerian Hospital where 50.2% were within the ages of 35-44 years.[3] but not in tandem with another study on SFA in west Ethiopia that reported more (45.8%) of their participants to be within 25-30 age bracket. [4]

Overall, this study findings revealed that about 9 out of 10 participants had at least one abnormality in their Seminal Fluid Analysis (SFA), which is similar to the findings of [3] where about four-fifth of the respondents were reported to have at least one abnormality in their SFA. Further analysis of the participants sperm count revealed 71.1% to have normozoospermia which is in tandem with [4] study on SFA analysis, only 3.0% had Azoospermia which is dissimilar to other study among the same study population that reported about half (47.5%) to have had Azoospermia. [3]

The findings of qualitative analysis in the study assessed the causes male infertility, the majority referred to lifestyle as the causes, while its evident some lifestyles like smoking, alcohol consumption, as well medical conditions as causes of male infertility This was supported by the study of [5]. This findings is also similar to the report of a qualitative study on male fertility.[6]

Male infertility according to the participants may cause psychological, emotional, and social distress, they stated that male infertility may cause by excessive drinking, having multiple sexual partners that may expose men to STIs and which may eventually cause infertility. Also, men believe that once they can ejaculate or fathered a child before, they cannot be infertile. And their diagnosis of been infertile may lead to emotional and psychological issues, some may even attempt suicidal ideation. Virtually all the respondents believed that male infertility can be caused but medical conditions. However, some of the participants in the qualitative study, including a doctor believed that male infertility can be caused by spiritual force. This is in tandem with the submission made by [6] study on lifestyle causes of male infertility.

## Conclusions

The findings indicate a prevalence of abnormal seminal fluid parameters (low sperm count, motility, and morphology) among the participants. Lifestyle choices were commonly cited as contributors to infertility, with participants highlighting the detrimental effects of habits such as smoking, STI and alcohol consumption on reproductive health. Additionally, lifestyle factors, including habitual smoking, STI and alcohol consumption, were recognized as potential predictors of abnormality in SFA and male infertility.

## Data Availability

All data produced in the present study are available upon reasonable request to the authors

## Acknowledgements

The authors wish to acknowledge the respondents who gave their consent to be part of this study as well as the head of the facility centres for their permission during the course of this study.

## Ethical approval and consent to participate

Ethical approvals for this study were obtained from the Ethics and Research Committee of Uniosun Teaching Hospital. The Protocol number UTH/EC/2024/06/957. All ethical principles guiding the conduct of research such as informed consent, beneficence, non-maleficence, confidentiality, justice, autonomy etc were strictly taken into consideration. All information gathered were confidential and privacy and confidentiality of the respondents were guaranteed. All respondents were made to sign the informed consent before filling the questionnaire.

